# Multicenter validation of artificial intelligence software predicting large vessel occlusion using noncontrast brain CT

**DOI:** 10.1101/2024.07.01.24309790

**Authors:** Jong-Won Chung, Myungjae Lee, Sue Young Ha, Pyeong Eun Kim, Leonard Sunwoo, Nakhoon Kim, Kwang-Yeol Park, Kyu Sun Yum, Dong-Ick Shin, Hong-Kyun Park, Yong-Jin Cho, Keun-Sik Hong, Jae Guk Kim, Soo Joo Lee, Joon-Tae Kim, Woo-Keun Seo, Oh Young Bang, Gyeong-Moon Kim, Dongmin Kim, Hee- Joon Bae, Wi-Sun Ryu, Beom Joon Kim

## Abstract

**Background:** To validate JLK-CTL, an artificial intelligence (AI) software developed to predict large vessel occlusion (LVO) using non-contrast CT (NCCT) scans, and to investigate its clinical implications regarding both infarct volume and functional outcomes.

**Methods:** Between January-2021 and April-2023, a consecutive series of patients who concurrently underwent CT angiography and NCCT within 24 hours of last-known-well (LKW) were collected. LVO was confirmed through consensus among three experts reviewing CT angiography. Infarct volumes were quantified using diffusion-weighted imaging (DWI) conducted within seven days of the NCCT. The performance of the JLK-CTL was evaluated based on the area under the receiver operating characteristic curve (AUROC), as well as its sensitivity and specificity. The association of JLK-CTL LVO scores with infarct volumes and functional outcomes was assessed using Pearson correlation and logistic regression analyses, respectively.

**Results:** Of 1,391 screened patients, 774 (mean age 69.0 ± 13.6 years, 57.6% men) were included. The median time from LKW to NCCT was 3.1 hours (IQR 1.5–7.4), with 24.2% (n=187) presenting LVO. The JLK-CTL demonstrated AUROC of 0.832 (95% CI 0.804–0.858), with a sensitivity of 0.711 (95% CI 0.641–0.775) and a specificity of 0.830 (95% CI 0.797–0.859) at the predefined threshold. Incorporating the National Institute of Health Stroke Scale into the model increased the AUROC to 0.872 (95% CI 0.846–0.894; p<0.001). The LVO scores showed a significant correlation with infarct volumes on follow-up DWI (r=0.53; p<0.001). When JLK-CTL LVO scores were categorized based on observed frequency of LVO, the highest JLK-CTL LVO scores (51-100) group showed an independent association with unfavorable functional outcomes (adjusted odds ratio 9.48; 95% CI 3.98–22.55).

**Conclusion:** The performance of the AI software in predicting LVO was validated across multiple centers. This tool has the potential to assist physicians in optimizing stroke management workflows, especially in resource-limited settings.

## Introduction

Large vessel occlusion (LVO) accounts for up to 46% of ischemic stroke^1^ and is associated with unfavorable outcome after ischemic stroke. Rapid and accurate diagnosis of LVO in the emergency room is essential for prompt intervention to mitigate further brain damage. However, patients presented with mild neurological deficits or atypical stroke symptoms are often misdiagnosed as having stroke mimics,^2^ posing a significant challenge to physicians who initially encounter patients with neurological symptoms. Given the critical importance of early recognition of LVO for timely interventions and improved functional outcomes, there remains an unmet clinical need for an accurate and rapid LVO diagnosis.

While non-contrast computed tomography (NCCT) scans are highly accessible and offer rapid acquisition, accurately diagnosing LVO from these scans requires expertise in interpreting NCCT scans and clinical correlation, which is often scarce in limited resources settings. Recent advances in artificial intelligence (AI) have facilitated the development of several software packages predicting LVO using NCCT. In a recent study,^3^ we reported an AI software (JLK-CTL, JLK Inc., Seoul, Republic of Korea) that predicts LVO solely based on NCCT images by analyzing hemispheric differences in Hounsfield units, regional brain volume, and the presence of a clot sign in the middle cerebral artery (MCA).

In this multicenter study involving six comprehensive stroke centers, we validated JLK-CTL in patients who visited hospitals within 24 hours of their last known well (LKW). In addition, we compared predicted and observed frequencies of LVO and categorized JLK-CTL LVO scores into four groups, which may provide more interpretable and dependable results for physicians. Furthermore, we investigated the clinical implications of JLK-CTL LVO scores in terms of infarct-relevant steno- occlusion of the MCA, infarct volumes on follow-up diffusion-weighted imaging (DWI), and 3-month functional outcomes after ischemic stroke.

## Materials and Methods

### Study populations

This multicenter study is stemmed from a brain imaging substudy of the ongoing nationwide stroke registry, Clinical Research Collaboration for Stroke in Korea (CRCS-K), which has recruited over 160,000 patients with stroke for 20 years.^4^ We consecutively enrolled patients with ischemic stroke or transient ischemic attack who admitted within 7 days of symptom onset from April 2022 to April 2023 at five comprehensive stroke centers (Figure S1). To ensure the heterogeneity of the data, we additionally collected a consecutive series of patients between January 2021 to March 2022 from Samsung Medical Center, which did not participate in the CRCS-K stroke registry. Inclusion criteria were 1) aged 18 years or older, and 2) concurrently underwent NCCT and CT angiography (CTA). Exclusion criteria were 1) NCCT performed beyond 24 hours of LKW, 2) poor image quality or insufficient contrast to analyze, 3) cases with hemorrhagic transformation, brain tumor, or external ventricular drain and 4) NCCT or CTA acquired after endovascular recanalization treatment. All patients or their legal representatives gave written informed consent. The study protocol was approved by the institutional review board of Seoul National University Bundang Hospital [B-2307-841-303].

### Clinical data collection

We retrieved baseline demographic and clinical information for all study participants from a web-based prospective stroke cohort (strokedb.or.kr).^5^ This included age, sex, history of previous stroke, functional status before stroke, and cardiovascular risk factors such as hypertension, diabetes mellitus, and atrial fibrillation. Stroke characteristics included the time interval between LKW and NCCT scan, the National Institutes of Health Stroke Scale (NIHSS) score at admission, and treatment information. Functional status at 3 months post-stroke was measured using the modified Rankin Scale (mRS) score, determined through a structured telephone interview by an experienced physician assistant at each hospital as previously described.^6,7^

### CT imaging protocols and analysis

NCCT images were acquired according to standard departmental protocols in each hospital. Section thickness ranged 3 ∼ 5 mm (Table S1). In the present study, LVO was defined as an arterial occlusion encompassing the intracranial segment of the internal carotid artery (ICA), as well as the M1 and M2 segments of the MCA (MCA-M1 and MCA-M2, respectively).^8^ To confirm the presence of LVO, CTA source images, maximum intensity projection images, and three-dimensional rendering images that were concurrently performed with NCCT were thoroughly examined by two experienced vascular neurologists (W-S. R and S.H), alongside an evaluation of patients’ follow-up magnetic resonance imaging (MRI) scans and symptomatic data. In cases of diagnostic discrepancy, a final determination was made by an experienced neuroradiologist (L. S). Along with the presence of LVO, location (ICA, MCA-M1, and MCA-M2) and the laterality of LVO were recorded. If the diagnosis was correct but the laterality of the LVO was discordant between JLK-CTL and the experts’ consensus, we designated the case as a false negative. We defined acute LVO as ICA or MCA-M1 occlusion relevant to the index stroke, whereas chronic LVO is defined as LVO irrelevant to the index stroke or LVO with hemodynamic infarct.^9,10^ Isolated MCA-M2 occlusion was defined as the presence of MCA-M2 occlusion without concurrent ICA or MCA-M1 occlusion. Infarct-relevant MCA stenosis is defined as moderate to severe stenosis on CTA that is related to infarcts observed on DWIs. For each NCCT scan, an experienced neurologist (J-W. C) rated Alberta Stroke Program Early CT score (ASPECTS).^11,12^

### Artificial intelligence software

NCCT images were processed through the validated and commercially available AI software (JLK-CTL, JLK Inc., Seoul, Korea) to predict LVO.^3^ In brief, the software analyzed differences in volume, tissue density, and Hounsfield unit distribution between bihemispheric regions (striatocapsular, insula, M1–M3, and M4–M6, modified from the ASPECTS). Additionally, the deep learning algorithm also automatically segmented hyperdense MCA signs as an extra feature. An ExtraTrees machine learning algorithm was employed to predict the JLK-CTL LVO score based on these features, which represents the likelihood of LVO determined by the algorithm. Furthermore, the software generated the JLK-CTL+ LVO score, incorporating NIHSS scores as an additional feature within the model.

### Follow-up imaging analysis

Follow-up DWI within 7 days after NCCT were included to analyze the association between JLK-CTL LVO score and follow-up infarct volumes. If the patient underwent two or more DWIs, we utilized the first image. Infarct volumes on DWI were calculated using a validated software package (JLK-DWI, JLK Inc., Seoul, Korea).^13–15^ The segmentation of the infarct area was carefully overseen by an experienced vascular neurologist (J-W. C). In cases where automated segmentation was inaccurate, manual corrections were applied to ensure precise segmentation.

### Categorization of LVO scores

We divided patients into deciles based on their JLK-CTL LVO scores, with each decile representing 10% intervals. For each decile, we calculated the observed frequency of LVO determined by expert consensus. Following this, we categorized the patients into four groups based on the observed frequency of LVO and the number of patients in each decile.

### Statistical analysis

Baseline characteristics among participating centers were compared using ANOVA or the Kruskal-Wallis test for continuous variables, and the chi-square test for categorical variables, as appropriate. To validate the accuracy of the JLK-CTL software in predicting LVO, we calculated the area under the receiver operating characteristic curve (AUROC), along with sensitivity, specificity, positive predictive value (PPV), and negative predictive value (NPV). We used a 1000-repeat bootstrap method to determine 95% confidence intervals (CIs) and compared AUROCs using the DeLong method.^16^ The cutoff for the JLK-CTL LVO score was set at 12.0 based on prior research.^3^ Additionally, we conducted the AUROC analysis after stratifying patients by participating centers.

After stratifying patients into five subgroups—acute LVO, chronic LVO, isolated MCA-M2 occlusion, infarct-relevant MCA stenosis, and no steno-occlusion of the MCA—we compared LVO scores using ANOVA with Tukey’s multiple comparison test. The association between JLK-CTL LVO scores and infarct volumes on DWI was analyzed using Pearson correlation analysis. Furthermore, we analyzed the association between JLK-CTL LVO scores and 3-month mRS scores using ANOVA and multivariable ordinal logistic and binary (mRS score 0–2 vs. 3–6) logistic regression analyses. Based on prior literature on functional outcomes after ischemic stroke,^7,17–19^ age, sex, admission NIHSS scores, previous stroke, hypertension, diabetes, atrial fibrillation, revascularization therapy, and time from LKW to NCCT scan were used as covariates. All statistical analyses were performed using STATA software (version 16.0, TX, USA) and MedCalc (version 17.2, MedCalc Software, Ostend, Belgium). A P value < 0.05 was considered statistically significant.

## Results

### Baseline characteristics

Among the 1,391 patients, 957 underwent concurrent NCCT and CTA. After excluding 183 patients, we included 774 patients in our analyses (Figure S1). The mean age of the included patients was 69.0 years (SD 13.6), and 446 (57.6%) were male. The median interval between LKW to NCCT scan was 3.1 hours (interquartile range 1.5–7.4) and 187 patients (24.2%) had LVO. Demographic and risk factor profiles were generally similar across participating centers, except for the prior history of stroke (Table 1). However, there were significant differences among the centers in terms of imaging vendors, time from LKW to NCCT scan, frequency of revascularization therapy, and the interval between NCCT and DWI (Table 1 and Table S1).

**Table 1.** Baseline characteristics of study population.

|  | Hospital A<br>(n = 116) | Hospital B<br>(n = 127) | Hospital C<br>(n = 96) | Hospital D<br>(n = 84) | Hospital E<br>(n = 104) | Hospital F<br>(n = 247) | P value |
| --- | --- | --- | --- | --- | --- | --- | --- |
| Age | 69.2±12.9 | 69.0±13.8 | 69.1±14.0 | 71.2±13.5 | 67.8±15.4 | 68.7±12.8 | 0.66 |
| Sex, male | 60 (51.7%) | 73 (57.5%) | 52 (54.2%) | 45 (53.6%) | 65 (62.5%) | 151 (61.1%) | 0.42 |
| Large vessel occlusion | 26 (22.4%) | 37 (29.1%) | 23 (24.0%) | 21 (25.0%) | 20 (19.2%) | 60 (24.3%) | 0.65 |
| Isolated MCA M2 occlusion | 6 (5.2%) | 6 (4.7%) | 1 (1.0%) | 5 (6.0%) | 5 (4.8%) | 16 (6.5%) | 0.48 |
| Initial NIHSS score | 3 (0 – 7) | 3 (1 – 9) | 3 (1 – 8) | 5 (1 – 10) | 4 (1 – 9) | 3 (1 – 8) | 0.16<sup>a</sup> |
| Previous stroke | 15 (12.9%) | 15 (11.8%) | 23 (24.0%) | 21 (25.0%) | 15 (14.4%) | 56 (22.7%) | 0.01 |
| Hypertension | 70 (60.3%) | 77 (60.6%) | 66 (68.7%) | 59 (70.2%) | 77 (74.0%) | 151 (61.1%) | 0.11 |
| Diabetes | 28 (24.1%) | 40 (31.5%) | 32 (33.3%) | 22 (26.2%) | 36 (34.6%) | 85 (34.4%) | 0.35 |
| Atrial fibrillation | 27 (23.3%) | 28 (22.0%) | 23 (24.0%) | 26 (31.0%) | 27 (26.0%) | 46 (18.6%) | 0.28 |
| High-risk cardioembolic source | 29 (25.0%) | 30 (23.6%) | 23 (24.0%) | 24 (28.6%) | 28 (26.9%) | 59 (24.0%) | 0.95 |
| CT vendor |  |  |  |  |  |  | < 0.001 |
| Philips | 116 (100.0%) | 3 (2.4%) | 0 | 0 | 0 | 0 |  |
| GE medical systems | 0 | 124 (97.6%) | 0 | 0 | 0 | 250 (99.6%) |  |
| SIEMENS | 0 | 0 | 96 (100.0%) | 81 (96.4%) | 0 | 0 |  |
| Toshiba | 0 | 0 | 0 | 3 (3.6%) | 104 (100.0%) | 1 (0.4%) |  |
| LKW to CT, hr | 3.5 (1.4 – 10.8) | 2.1 (1.2 – 4.2) | 5.3 (2.3 – 9.4) | 1.6 (1.1 – 2.5) | 2.9 (1.6 – 6.3) | 4.8 (2.2 – 9.8) | < 0.001<sup>a</sup> |
| Revascularization therapy |  |  |  |  |  |  | < 0.001 |
| Intravenous only | 5 (4.3%) | 14 (11.0%) | 11 (11.5%) | 12 (14.3%) | 14 (13.5%) | 11 (4.6%) |  |
| Endovascular therapy only | 11 (9.5%) | 15 (11.8%) | 5 (5.2%) | 13 (15.5%) | 6 (5.8%) | 27 (11.2%) |  |
| Combined | 12 (10.3%) | 18 (14.2%) | 4 (4.2%) | 5 (6.0%) | 5 (4.8%) | 6 (2.5%) |  |
| Interval between NCCT and DWI, hr | 2.2 (1.2 – 3.9) | 0.5 (0.2 – 2.1) | 0.8 (0.4 – 2.5) | 1.9 (1.3 – 3.9) | 2.1 (1.3 – 7.8) | 1.8 (1.3 – 2.8) | < 0.001<sup>a</sup> |
| Infarct volume on DWI, mL | 2.8 (0.3 – 22.7) | 1.8 (0.3 – 11.0) | 3.4 (1.1 – 32.1) | 0.5 (0.1 – 3.2) | 0.6 (0.1 – 6.2) | 0.9 (0.2 – 12.2) | < 0.001<sup>a</sup> |

Data are presented as mean ± standard deviation, number (percentage), and median (interquartile range).
<sup>a</sup>Kruskal-Wallis test was used. MCA=middle cerebral artery; NIHSS=National Institute Health Stroke Scale; LKW=last known well; NCCT=noncontrast CT; DWI=diffusion-weighted imaging.

### Efficacy of JLK-CTL in multicenter dataset

Overall, JLK-CTL achieved an AUROC of 0.832 (95% CI 0.804–0.858), which was significantly higher than the AUROC obtained using the ASPECTS (0.755 [95% CI 0.723–0.785]; p<0.001) and comparable to the AUROC obtained using NIHSS scores (0.837 [95% CI 0.809–0.862]; p=0.82; Figure 1). At the predetermined JLK-CTL LVO score threshold of 12.0, JLK-CTL demonstrated a sensitivity of 0.711 (95% CI 0.641–0.775), specificity of 0.830 (95% CI 0.797–0.859), PPV of 0.571 (95% CI 0.505–0.635), and NPV of 0.900 (95% CI 0.872–0.924; Table 2).

**Figure 1.**
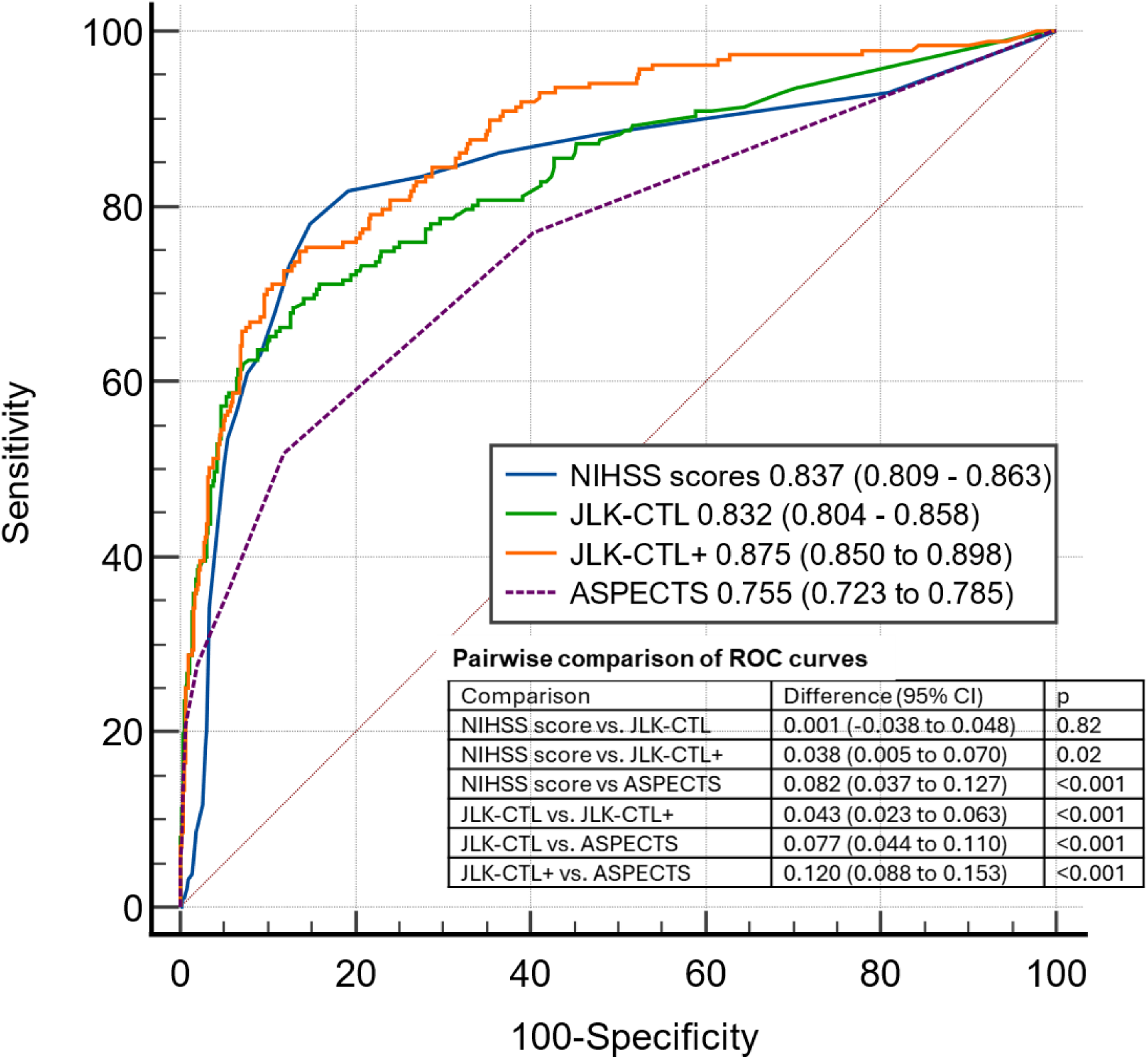
Receiver operating characteristic curves comparing NIHSS scores, JLK- CTL, JLK-CTL+, and ASPECTS for predicting large vessel occlusion using noncontrast brain CT scans. NIHSS=National Institute Health Stroke Scale; ASPECTS= Alberta stroke program early CT score; CI=confidence interval

**Table 2.** Performance of artificial Intelligence software in the entire population and subgroups stratified by time from last- known-well to noncontrast CT scans (< 6 hours vs. 6-24 hours)

| Confusion matrix | All patients (N = 774) |  | LKW to NCCT < 6 hours (n = 537) |  | LKW to NCCT > 6 and ≤ 24 hours (n = 231) |  |
| --- | --- | --- | --- | --- | --- | --- |
|  | Prediction |  | Prediction |  | Prediction |  |
|  | LVO | No LVO | LVO | No LVO | LVO | No LVO |
| Ground truth, LVO | 133 | 54 | 108 | 40 | 24 | 13 |
| Ground truth, no LVO | 100 | 487 | 71 | 318 | 28 | 166 |
| Sensitivity (95% CI) | 0.711 (0.641 – 0.775) |  | 0.730 (0.651 – 0.799) |  | 0.649 (0.475 – 0.798) |  |
| Specificity (95% CI) | 0.830 (0.797 – 0.859) |  | 0.817 (0.775 – 0.855) |  | 0.856 (0.798 – 0.902) |  |
| PPV (95% CI) | 0.571 (0.505 – 0.635) |  | 0.603 (0.528 – 0.676) |  | 0.462 (0.322 – 0.605) |  |
| NPV (95% CI) | 0.900 (0.872 – 0.924) |  | 0.888 (0.851 – 0.919) |  | 0.927 (0.879 – 0.961) |  |
| AUROC | 0.832 (0.804 – 0.858) |  | 0.839 (0.805 – 0.869) |  | 0.796 (0.738 – 0.846) |  |

LKW=last-known-well; NCCT=noncontrast CT; LVO=large vessel occlusion; PPV=positive predictive value; NPV=negative predictive value; AUROC=area under the receiver operating characteristics curve.

When stratified by time from LKW to NCCT scans (< 6 hours vs. 6–24 hours), JLK-CTL exhibited higher sensitivity in the < 6 hours group while higher specificity in the 6-24 hours group. Incorporating the NIHSS score (JLK-CTL+) increased the AUROC to 0.872 (95% CI 0.846–0.894), significantly outperforming both JLK-CTL and NIHSS scores alone (p<0.001 and p=0.035, respectively). JLK-CTL+ demonstrated a sensitivity of 0.733 (95% CI 0.663–0.795) and specificity of 0.847 (95% CI 0.815–0.875) in the entire study population (Table S2).

The performance of the JLK-CTL varied across participating centers, with AUROCs ranging from 0.754 to 0.927, sensitivity from 0.423 to 0.857, and specificity from 0.753 to 0.911 (Table 3). Excluding patients with chronic occlusion (n=30), JLK- CTL and JLK-CTL+ achieved AUROCs of 0.867 (95% CI 0.840–0.890) and 0.907 (95% CI 0.884–0.927), respectively (Figure S3). Sensitivity also increased to 0.761 (95% CI 0.688–0.824) and 0.791 (95% CI 0.721–0.851), respectively (Table S3).

**Table 3.** Performance of artificial Intelligence software in each participating center.

| Confusion matrix | Hospital A (n = 116) |  | Hospital B (n = 127) |  | Hospital C (n = 96) |  | Hospital D (n = 84) |  | Hospital E (n = 104) |  | Hospital F (n = 247) |  |
| --- | --- | --- | --- | --- | --- | --- | --- | --- | --- | --- | --- | --- |
|  | Prediction |  | Prediction |  | Prediction |  | Prediction |  | Prediction |  | Prediction |  |
|  | LVO | No LVO | LVO | No LVO | LVO | No LVO | LVO | No LVO | LVO | No LVO | LVO | No LVO |
| Ground truth, LVO | 11 | 15 | 27 | 10 | 18 | 5 | 18 | 3 | 14 | 6 | 45 | 15 |
| Ground truth, no LVO | 8 | 82 | 16 | 74 | 18 | 55 | 11 | 52 | 15 | 69 | 32 | 155 |
| Sensitivity (95% CI) | 0.423 (0.234 – 0.631) |  | 0.730 (0.559 – 0.862) |  | 0.783 (0.563 – 0.925) |  | 0.857 (0.637 – 0.970) |  | 0.700 (0.457 – 0.881) |  | 0.750 (0.621 – 0.853) |  |
| Specificity (95% CI) | 0.911 (0.832 – 0.961) |  | 0.822 (0.727 – 0.895) |  | 0.753 (0.639 – 0.847) |  | 0.825 (0.709 – 0.909) |  | 0.821 (0.723 – 0.896) |  | 0.829 (0.767 – 0.880) |  |
| PPV (95% CI) | 0.579 (0.335 – 0.797) |  | 0.628 (0.467 – 0.770) |  | 0.500 (0.329 – 0.671) |  | 0.621 (0.423 – 0.793) |  | 0.483 (0.294 – 0.675) |  | 0.584 (0.466 – 0.696) |  |
| NPV (95% CI) | 0.845 (0.758 – 0.911) |  | 0.881 (0.792 – 0.941) |  | 0.917 (0.816 – 0.972) |  | 0.945 (0.849 – 0.989) |  | 0.920 (0.834 – 0.970) |  | 0.912 (0.859 – 0.950) |  |
| AUROC | 0.754 (0.665 – 0.829) |  | 0.809 (0.729 – 0.873) |  | 0.843 (0.755 – 0.909) |  | 0.927 (0.850 – 0.973) |  | 0.839 (0.754 – 0.904) |  | 0.847 (0.796 – 0.889) |  |

LVO=large vessel occlusion; PPV=positive predictive value; NPV=negative predictive value; AUROC=area under the receiver operating characteristics curve.

### Associations of LVO scores with steno-occlusion of middle cerebral artery and infarct volumes on follow-up diffusion-weighted imaging

In comparison to other groups, the acute LVO group (n=126) exhibited a higher JLK- CTL LVO score, with a median of 34.4 (IQR 14.6–57.0; Figure 2). In addition, the isolated MCA-M2 occlusion group (n=38) had higher LVO scores (median 16.1; IQR 8.0–33.5) compared to the group with infarct-relevant MCA stenosis (n=36; median 8.0; IQR 7.8–10.3) and those without steno-occlusion of the MCA (n=551; median 7.8; IQR 7.6–9.3). DWIs were available for 745 patients with a median interval of 1.7 hours (IQR 0.9–3.2) from the NCCT scan. JLK-CTL LVO scores were significantly correlated with infarct volumes on follow-up DWI (r=0.53; p<0.001; Figure 3A). This correlation was stronger in patients with LVO (n=181; r=0.47; Figure 3B) compared to those without LVO (n=564; r=0.16; Figure 3C).

**Figure 2.**
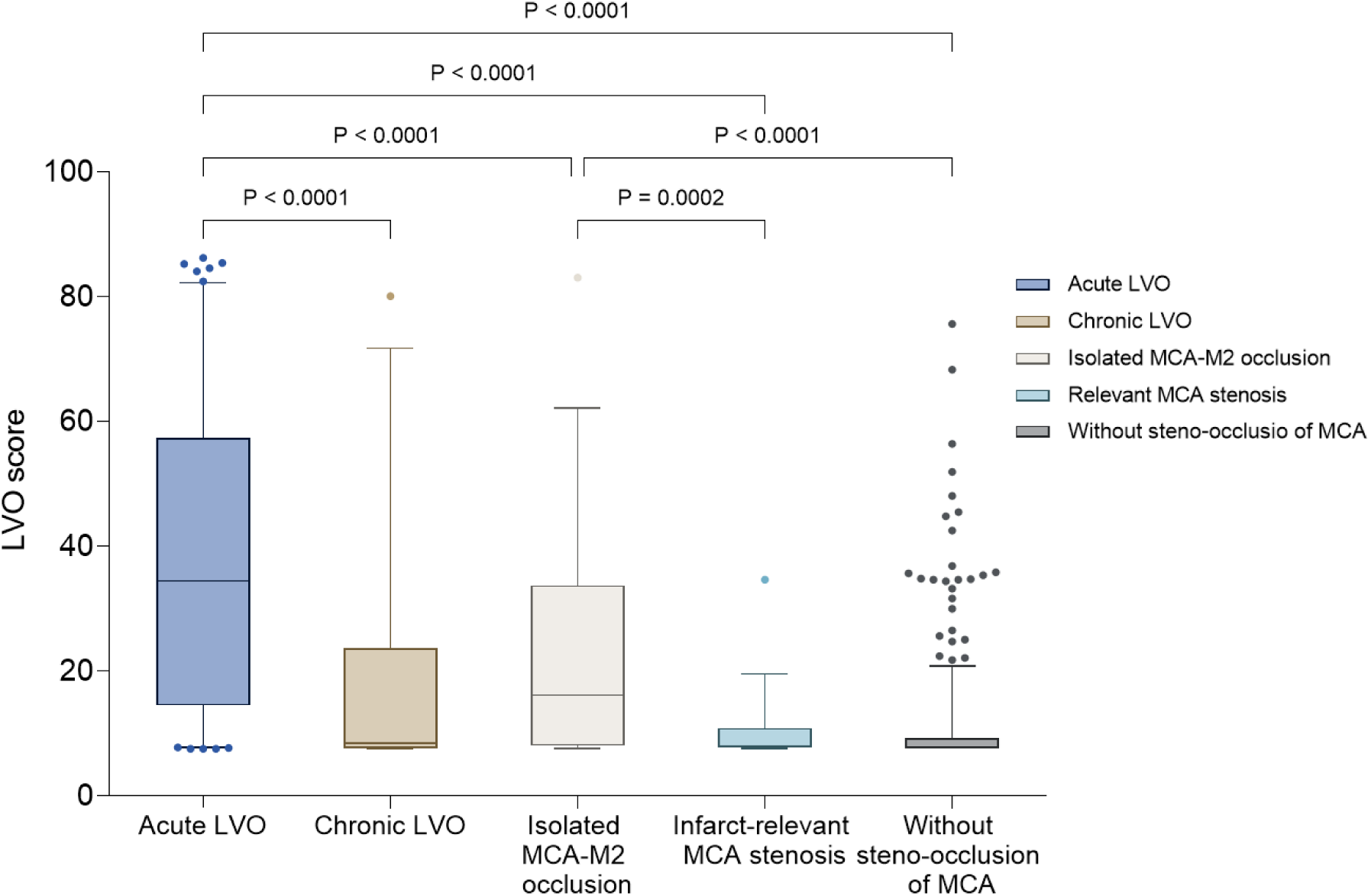
Box plots illustrating JLK-CTL LVO scores across subgroups categorized by steno-occlusion of the middle cerebral artery (MCA). Statistical comparisons were conducted using ANOVA followed by Tukey post-hoc analysis. LVO=large vessel occlusion.

**Figure 3.**
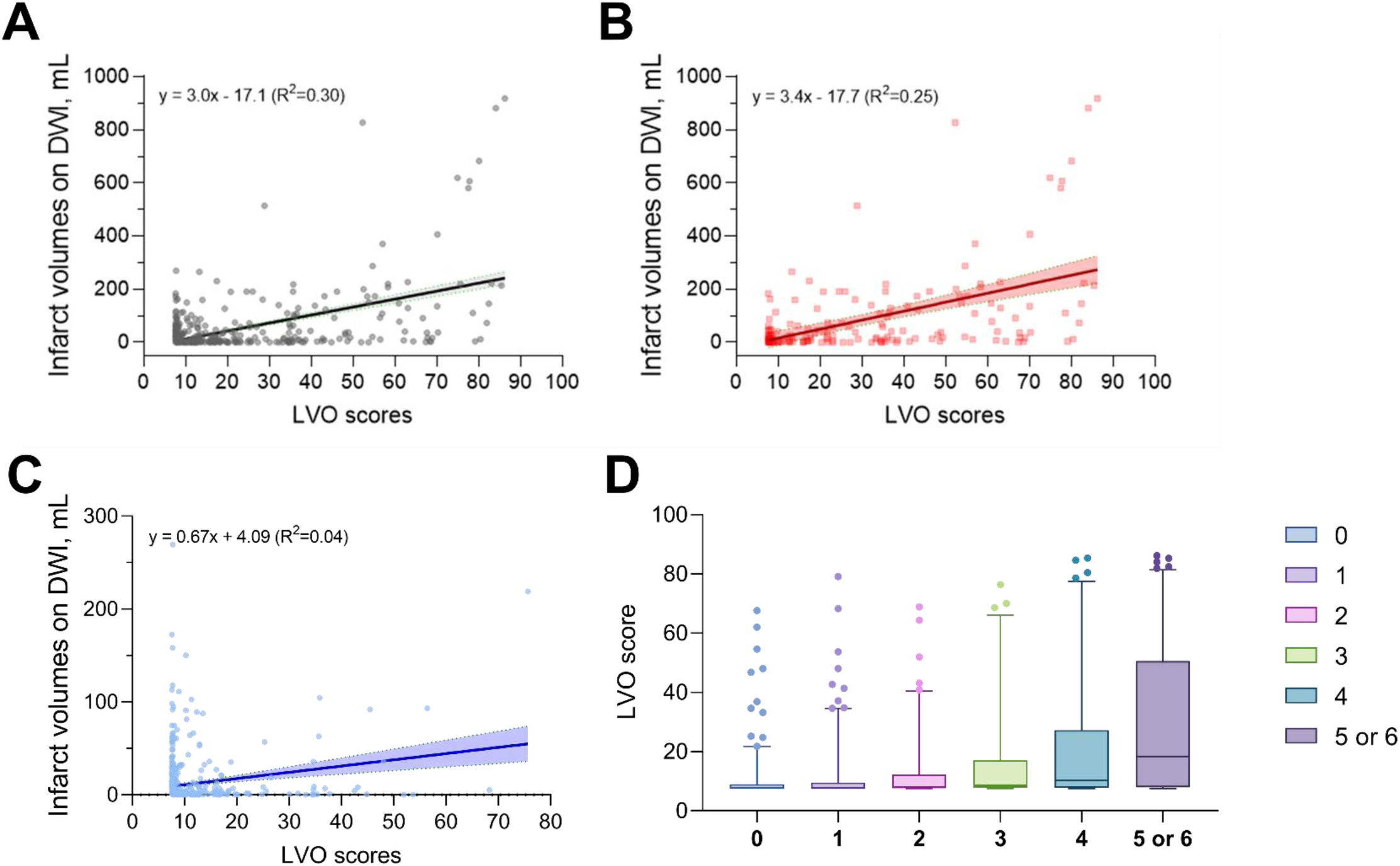
Dot plots illustrate the relationship between JLK-CTL LVO scores and infarct volumes on follow-up diffusion-weighted imaging (DWI) in (A) all patients, (B) patients with LVO, and (C) patients without LVO. The black, red and blue lines with shaded areas represent linear regression lines and their 95% confidence intervals for patients with and without large vessel occlusion (LVO), respectively. (D) Box plot showing JLK-CTL LVO scores stratified by modified Rankin Scale score at 3 months after ischemic stroke.

### Categorizing JLK-CTL LVO scores using observed frequency of LVO in multicenter data

LVO scores demonstrated a right-skewed distribution (Figure S4). When patients were stratified into deciles based on their LVO scores, a nearly linear increase in LVO frequency was observed with higher scores (Figure 4A). However, due to the small number of patients in groups with LVO scores > 40, interpretations from these groups may be less reliable. Therefore, we categorized LVO scores into four groups: 0-10 (unlikely), 11-20 (less likely), 21-50 (possible), and 51-100 (suggestive), considering both the patient distribution and observed LVO frequencies. After categorization, the observed frequencies of LVO were 9.8%, 23.1%, 71.6%, and 91.7% in unlikely, less likely, possible and suggestive groups, respectively (Figure 4B).

**Figure 4.**
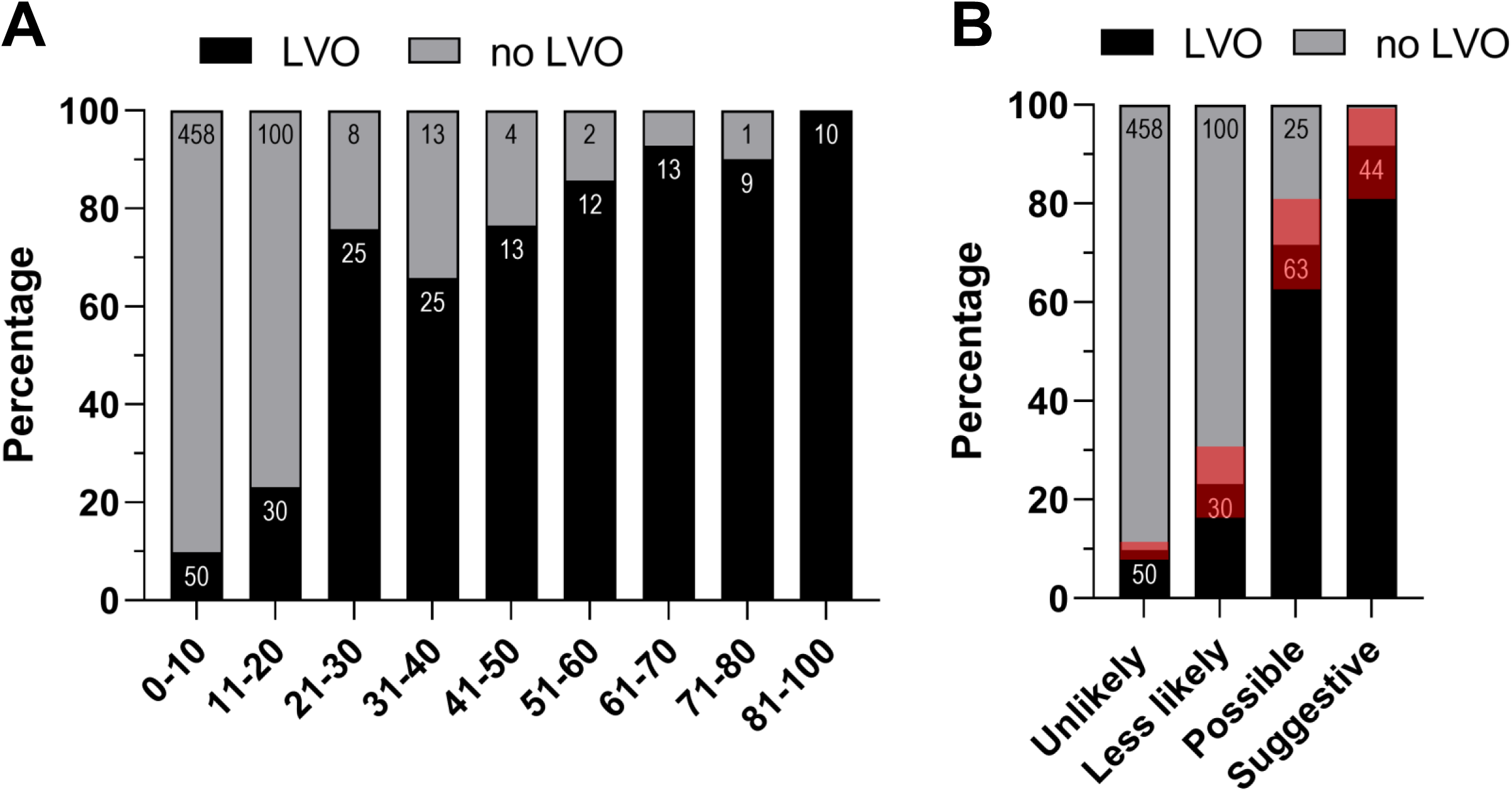
(A) Observed frequency of large vessel occlusion (LVO) across deciles of JLK-CTL LVO scores. Numbers within bars indicate the patient count. (B) Observed frequency of LVO across groups categorized as unlikely (LVO score 0–10), less likely (11–20), possible (21–50), and suggestive (50–100) based on observed frequencies of LVO. Red boxes represent the 95% confidence intervals for the observed frequency of LVO.

### Continuous or categorized JLK-CTL LVO scores and functional outcomes

JLK-CTL LVO scores were significantly associated with 3-month mRS scores after ischemic stroke (p<0.001; Figure 3D). Multivariable ordinal and binary logistic regression analyses confirmed an independent relationship between LVO scores and 3-month functional outcomes after ischemic stroke (Table 4 and Table S4). After adjusting for covariates, each 1-point increase in the LVO score was associated with a 2% higher likelihood of worse mRS scores or unfavorable outcomes. Additionally, possible (JLK-CTL LVO score 21–50) and suggestive (51–100) LVO groups were independently associated with higher mRS scores, with adjusted odds ratios of 1.96 (95% CI 1.25–3.07) and 2.35 (95% CI 1.30–4.26), respectively. These groups were also associated with unfavorable functional outcomes, with adjusted odds ratios of 2.57 (95% CI 1.45–4.54) and 9.48 (95% CI 3.98–22.55), respectively.

**Tabe 4.** Multivariable ordinal and binary logistic regression analysis between 3-month functional outcome and LVO scores as a continuous or categorical variable.

|  | Unadjusted | p | Adjusted* | p |
| --- | --- | --- | --- | --- |
|  | Odds ratio |  | Odds ratio |  |
| Ordinal logistic |  |  |  |  |
| LVO score, per 1 score | 1.04 (1.03 – 1.05) | < 0.001 | 1.02 (1.01 – 1.03) | < 0.001 |
| Categorized LVO score |  |  |  |  |
| 0 – 10 (n = 508) | Reference |  | Reference |  |
| 10 – 20 (n = 130) | 1.70 (1.19 – 2.41) | 0.003 | 1.07 (0.74 – 1.53) | 0.73 |
| 20 – 50 (n = 88) | 4.91 (3.25 – 7.43) | < 0.001 | 1.96 (1.25 – 3.07) | 0.003 |
| 50 – 100 (n = 48) | 10.86 (6.34 – 18.60) | < 0.001 | 2.35 (1.30 – 4.26) | 0.005 |
| Binary logistic |  |  |  |  |
| LVO score, per 1 score | 1.05 (1.04 – 1.06) | < 0.001 | 1.02 (1.00 – 1.03) | 0.039 |
| Categorized LVO score |  |  |  |  |
| 0 – 10 (n = 508) | Reference |  | Reference |  |
| 10 – 20 (n = 130) | 1.84 (1.21 – 2.80) | 0.004 | 1.28 (0.79 – 2.07) | 0.31 |
| 20 – 50 (n = 88) | 4.10 (2.53 – 6.66) | < 0.001 | 2.57 (1.45 – 4.54) | 0.001 |
| 50 – 100 (n = 48) | 14.83 (6.74 – 32.66) | < 0.001 | 9.48 (3.98 – 22.55) | < 0.001 |

\*Adjusted for age, sex, admission NIHSS scores, previous stroke, hypertension, diabetes, atrial fibrillation, revascularization therapy, and time from LKW to NCCT scan

## Discussion

In this multicenter study utilizing data from a nationwide registry, we validated JLK- CTL, a software designed to predict LVO on NCCT. The JLK-CTL achieved a sensitivity of 0.711 and a specificity of 0.830 at the predefined threshold, with performance varying among hospitals. JLK-CTL LVO scores correlated with infarct volumes on follow-up DWI and 3-month mRS scores. Based on the observed frequency of LVO, we categorized JLK-CTL LVO scores into four groups, which accurately represented the observed frequency of LVO and were independently associated with functional outcomes after ischemic stroke.

Consistent with our previous findings,^3^ the JLK-CTL reliably predicted LVO on NCCT. JLK-CTL outperformed the ASPECTS and showed comparable performance to the NIHSS score, suggesting that JLK-CTL could streamline stroke management workflows, especially for less experienced physicians. Moreover, integrating NIHSS scores into the algorithm increased the sensitivity, enabling detection of three-quarters of LVO cases with high specificity. This capability has the potential to assist stroke specialists in promptly initiating treatment for patients with LVO.

During our evaluation of JLK-CTL at each participating center, we observed varying performance metrics, with AUROC ranging from 0.754 to 0.927. Despite similar baseline characteristics among stroke centers and no significant associations found between time from LKW to NCCT scan or revascularization rates and JLK-CTL performance, we hypothesize that inherent overfitting of the AI model may explain these discrepancies.^20,21^ Since our algorithm relies primarily on Hounsfield unit differences between hemispheres as a crucial feature, inter-scanner variability in Hounsfield units^22–24^ could potentially contribute to the observed variability in JLK- CTL performance.

In the present study, we observed a notably skewed distribution of JLK-CTL LVO scores, posing challenges for calibration in ranges with limited data. Moreover, the varying frequency of LVO across participating centers suggests that a model-based calibration, commonly used in deep learning algorithms,^25^ is less practical and prone to miscalibration due to the highly variable disease prevalence, diverse scanners, and varying imaging parameters in clinical settings. To address these challenges, we consolidated multiple categories with similar observed frequencies of LVO into four distinct groups. This approach aimed to provide a more reliable interpretation of the data that better reflects the actual occurrence of LVO across our study cohort. We believe that this calibrated categorization enhances the clinical utility of our findings for physicians and healthcare providers.

We found that patients with isolated MCA-M2 occlusion had significantly higher JLK-CTL LVO scores compared to those with infarct-relevant MCA stenosis or without steno-occlusion of the MCA. This finding extends our previous study,^3^ which was restricted to proximal MCA occlusion, to include more distal MCA occlusions. Since the algorithm utilizes ASPECTS regions M1-M3 and M4-M6 as radiomics features, it was able to detect isolated MCA-M2 occlusions, although their LVO scores were lower than those for acute proximal LVO.

ASPECTS is negatively associated with infarct volume on DWI.^26^ However, inconsistent results have been reported regarding the association between ASPECTS on baseline NCCT and functional outcomes.^27,28^ Despite a very short time interval between LKW to NCCT scans (median 3.1 hours) in the present study, we observed a significant correlation between the JLK-CTL LVO score and infarct volume on DWI which was obtained with a median interval of 1.7 hours between NCCT scans and DWI. In addition, we found that both JLK-CTL LVO score and the categorized LVO score predict unfavorable outcomes after ischemic stroke. Specifically, patients with an JLK- CTL LVO score greater than 50 had a 9.5-fold increased risk of unfavorable outcomes compared to those with an JLK-CTL LVO score of 0-10, after adjusting for covariates. If warranted in future studies, the JLK-CTL LVO score could become a clinically important radiomic marker for patients with ischemic stroke.

Our study has limitations. First, we included only an Asian stroke population. The etiology of LVO differs between ethnicities,^29,30^ which could potentially affect the software’s performance. Future studies should include multinational, multiethnic populations to validate the algorithm’s generalizability. Second, we only tested JLK- CTL in patients diagnosed with ischemic stroke. As a result, the clinical efficacy, resource utilization, and cost-effectiveness of the software were not fully investigated. Third, JLK-CTL can detect only ICA or MCA occlusion, and thus is not applicable to anterior cerebral artery, posterior cerebral artery, and posterior circulation LVO.

In conclusion, we demonstrated the clinical efficacy of JLK-CTL in predicting LVO on NCCT using a multicenter dataset. Additionally, JLK-CTL LVO scores correlated with infarct volumes on follow-up DWI and 3-month functional outcomes after ischemic stroke. The software may assist physicians in rapidly identifying stroke patients who require further investigation and treatment.

## Data Availability

The anonymized data used for this study will be made available upon reasonable request and the approval of our Data Steering Committee.

## Acknowledgment

The authors appreciate the contributions of all members of the Clinical Research Collaboration for Stroke in Korea to this study.

## Supplemental Material

Table S1–S4 Figure S1–S4

## Non-standard Abbreviations and Acronyms

LVO: large vessel occlusion
NCCT: Noncontrast computed tomography AI Artificial Intelligence
LKW: Last known well
CTA: computed tomography angiography NIHSS National Institutes of Health Stroke Scale
mRS: modified Rankin Scale
ICA: internal carotid artery MCA middle cerebral artery
ASPECTS: Alberta Stroke Program Early CT Score
AUROC: area under the receiver operating characteristic curve PPV positive predictive value
NPV: negative predictive value

